# Routine discussion of previous trauma in the perinatal period: Insights from interviews with women, voluntary sector representatives, and maternity care providers

**DOI:** 10.1101/2024.10.01.24314700

**Authors:** Joanne Cull, Gill Thomson, Soo Downe, Michelle Fine, Anastasia Topalidou

## Abstract

**Background:** Many pregnant women have a history of trauma, such as abuse or violence, which can significantly impact their mental and physical health. Discussing these experiences in maternity care presents an opportunity to support women, reduce stigma, and connect them with resources. However, concerns persist about stigmatisation, re-traumatisation and unwarranted safeguarding referrals. As part of a larger study that aimed to develop a methodology for conducting trauma discussions, interviews were carried out with a range of stakeholders.

**Methods:** Semi-structured interviews were conducted with women with trauma histories (experts by experience; n=4), representatives of voluntary sector organisations (n=7), and healthcare providers (n=12). Reflexive thematic analysis was used to analyse the data. The study employed a critical participatory action research approach, supported by a Patient and Public Involvement & Engagement group (named as the ‘Research Collective’ for this study) comprising experts by experience, maternity care professionals, and voluntary sector practitioners. The group contributed to both the design and analysis phases of the research.

**Findings:** Five key themes emerged from the interviews, exploring both the benefits and challenges of trauma discussion in maternity care. Participants reflected on who should lead these discussions, the appropriate settings and timings, and strategies for effective communication. The emotional and training needs of care providers conducting trauma discussions were also highlighted.

**Conclusion:** Trauma discussions in maternity care are a complex but necessary intervention that require careful consideration of timing, communication, and referral pathways. This paper offers concrete steps towards creating a more empathetic and supportive maternity care environment.

**Statement of Significance:** *Problem:* Traumatic experiences such as abuse or violence contribute to long term mental and physical health problems.

*What is already known:* Raising the issue of previous trauma within maternity care offers an opportunity to provide support, but if handled insensitively can be distressing to women.

*What this paper adds:* This paper shows that discussing trauma is complex and requires a system-wide approach which addresses when, where, and how to talk about trauma, referral pathways, and the need for training and support for maternity care providers. It also offers insights on conducting these discussions sensitively and effectively.

## Introduction

Many pregnant women have histories of violence, trauma and abuse. For example, in the United Kingdom, a quarter of women have suffered physical, sexual, or emotional abuse or witnessed domestic abuse before the age of 16 while 59% of young women in Australia report at least one adverse childhood experience (Office for National Statistics, 2020; Loxton et al., 2021). These experiences can have profound impacts for mental and physical health, as well as health-seeking behaviours, with long-lasting consequences (Bellis et al., 2017; Bellis et al., 2019; Hughes et al., 2017). Women who have suffered previous trauma face heightened risk of relapses of existing mental health conditions and the onset of new disorders during the perinatal period (Young-Wolff et al., 2019).

Discussing previous trauma within maternity care offers an opportunity to inform pregnant women about its potential impact and guide them towards relevant support such as substance abuse treatment or mental health services (Flanagan et al., 2018). Nonetheless, concerns persist regarding the potential for re-traumatisation, unwarranted safeguarding referrals, and stigmatisation of women with traumatic histories (Ford et al., 2019; Underwood, 2020; Racine, Killam & Madigan, 2020).

To inform the development of an intervention focused on routine trauma discussions in maternity care, we conducted interviews with women with trauma histories (referred to as experts by experience in this paper), representatives of voluntary sector organisations, and healthcare providers. In this context, we define ‘routine’ as raising the issue of previous trauma with all women accessing maternity services, rather than selectively for those suspected of having experienced trauma.

### Reflexive note

We explored our pre-existing beliefs on routine trauma discussions and their potential impact on the study. JC and SD, midwives and maternity care researchers, shared concerns about potential harm caused by poorly conducted trauma conversations. GT, a perinatal mental health researcher, advocated for trauma-informed conversations. AT, a maternal and neonatal care researcher, stressed the importance of supportive care models. MF, an expert in critical psychology and participatory research, saw trauma as a complex source of knowledge and creativity.

The study employed critical participatory action research methodology and was supported by a Patient and Public Involvement & Engagement group (named as the ‘Research Collective’ for this study) comprising experts by experience, maternity care professionals, and voluntary sector practitioners. Their involvement in framing the study, developing interview questions, selecting participants, and analysing data was crucial in challenging preconceived ideas. Regular team discussions and feedback from the Research Collective upheld trustworthiness, and anonymous feedback after workshops aimed to ensure all Collective members felt heard and able to contribute. Findings were shared at national and international conferences which gave opportunities for peer reflection. JC maintained reflexivity through journaling, supervision, and counselling, further enhancing analytical rigour.

### Study design

In accord with a critical participatory action research approach, the Research Collective played a crucial role in shaping the study methods and interview questions. Recognising the sensitivity of the topic and the importance of ensuring participant comfort and confidentiality, one-to-one interviews were chosen for data collection.

### Recruitment

Following discussions with the Research Collective, purposive sampling was undertaken to ensure representation from various maternity care professions, experience levels, and demographics, as well as diverse trauma types among voluntary sector practitioners and experts by experience.

Maternity care professionals and experts from the voluntary sector were recruited through the clinical networks of the research team and by approaching professionals and experts known to be working in this area. The recruitment of experts by experience was facilitated through collaborating with voluntary sector organisations dedicated to supporting women following previous trauma. These organisations distributed a recruitment flyer, inviting interested women to contact the research team direct. We aimed to respect participants’ autonomy by not requiring disclosure of personal experiences to establish eligibility. Eligibility criteria included being over 18 and having accessed UK maternity services at any previous time.

### Ethics

Ethical approval from the University of Central Lancashire Health Ethics Review Panel was obtained (reference HEALTH 0220). All participants received a £10 ‘thank you’ voucher.

Recruiting experts by experience through voluntary sector organisations ensured that women did not feel coerced into participating; it also ensured that participants had access to emotional support. All recruitment discussions occurred via email, further mitigating any perceived pressure. Prior to participation, all potential participants received comprehensive study documentation, including the interview topic guide, enabling individuals to fully consider their involvement before committing.

The interview topic guide was carefully designed with the Research Collective to minimise distress among participants. Participants were not questioned about their trauma histories or personal maternity care experiences. A distress protocol was developed in case a participant should become upset during an interview. The participant information sheet stated that the interviewee could choose not to answer any question, and that they were free to stop the interview at any time and without giving a reason. Participants were also made aware that confidentiality was assured unless they disclosed that they or others were at risk of harm. The information sheet provided details of available support services, and following the interview, participants received a debrief email reiterating those details. At the outset of the interview, participants were asked to either sign a consent form (for face-to-face interviews) or provide verbal consent (for Teams interviews).

To safeguard anonymity, each interviewee was assigned a unique code number, which was used to label all documents instead of their names. Certain demographic data were aggregated, and job titles were generalised to protect the identities of participants in unique or national roles.

### Data collection

As noted above, the Research Collective were actively engaged in shaping the interview topic guide (Appendix 1), providing invaluable insights into the content, sequence, and language of the questions. The guide included questions relevant to all participants, such as *‘When should maternity care providers inquire about difficult experiences?*’ Additionally, questions tailored specifically to experts from the voluntary sector and healthcare professionals were included, such *as ‘How can adequate time be allocated for these discussions?’* A selection of trauma screening tools, identified while carrying out a systematic review which preceded this study (Cull et al., 2023), were also used as prompts.

Two interviews were undertaken face to face, and the remainder (n=20, one of which had two participants) by Microsoft Teams. The interviews took an average of 60-90 minutes. A professional transcription service, with appropriate confidentiality agreements, was used to ensure accuracy. Recruitment was stopped after 23 participants, as rich data had been obtained and a range of diverse perspectives engaged.

### Data analysis

Reflexive thematic analysis, defined as ‘*a method for identifying, analysing and reporting patterns (themes) within data’* (Braun and Clarke, 2006, p.79) was employed to analyse the collected data. This approach allows for a nuanced exploration of participants’ views, facilitating the identification of underlying meanings and patterns within the data.

The thematic analysis followed Braun and Clarke’s six-step approach to qualitative data analysis (2006). In the initial phase of familiarisation, each transcript was thoroughly reviewed to gain a deep understanding of the data. During the generation of initial codes, the entire dataset was systematically examined for relevant data, and tentative codes were created. This process involved multiple iterations to ensure consistency and accuracy in coding. During the third phase of searching for themes, codes were reviewed and grouped into broader patterns of meaning, with attention given to outliers. Alignment with coded extracts and the overall dataset was then assessed, and themes were refined accordingly (fourth phase). In the fifth phase, themes were described and named, with input from the whole research team to enhance trustworthiness and mitigate personal biases. The findings were also shared and refined within two workshops with the Research Collective.

## Results

### Participants

Of the 23 participants who took part in this study, 12 were maternity care providers, seven were voluntary sector practitioners and four were experts by experience. A summary of participants’ demographic data can be found in Table 1, and pseudonymised job titles for maternity care professional participants in Table 2. Practitioners from across a range of relevant professions were represented, including midwifery, obstetrics, perinatal mental health, health visiting, psychosexual therapy, psychiatry, children’s social care and general practice. The voluntary sector practitioners specialised in supporting women after domestic abuse, birth trauma, removal of children from care, seeking asylum, sexual violence, and female genital mutilation.

**Table 1.** Participant demographic data.

|  | **Experts by experience (n=4)** | **Maternity care providers and experts from the voluntary sector (n=19)** |
| --- | --- | --- |
| **Sex** |  |  |
| Female | 4 | 17 |
| Male | 0 | 2 |
| **Self-described ethnic category** |  |  |
| White, White British or White Other | 3 | 15 |
| Black African or African Black British | 1 | 4 |
| **Age** |  |  |
| 18-30 | 1 | 0 |
| 31-45 | 3 | 9 |
| 46-60 | 0 | 6 |
| Over 60 | 0 | 4 |
| **Region** |  |  |
| England | 4 | 15 |
| Wales | 0 | 2 |
| Scotland | 0 | 2 |

**Table 2.** Job titles - maternity care professional participants (n=12)

|  | Job Title |
| --- | --- |
| 1. | Clinical matron / specialist midwife |
| 2. | Specialist midwife for perinatal mental health |
| 3. | Consultant Liaison Psychiatrist |
| 4. | Health Visitor |
| 5. | Clinical Lead for Perinatal Mental Health |
| 6. | Clinical Lead for Perinatal Mental Health in a prison |
| 7. | Team Manager, Children's Social Care |
| 8. | Professional Midwifery Advocate |
| 9. | General Practitioner (retired) |
| 10. | Psychosexual therapist |
| 11. | Midwife - community and hospital |
| 12. | Consultant Obstetrician and Gynaecologist |

The categorisation of participants in the study proved to be more nuanced and overlapping than initially anticipated. Many maternity care professionals and voluntary sector practitioners shared their own experiences of trauma or that of their family members during the interviews. One of the voluntary sector practitioners is a qualified midwife with a wealth of experience supporting women in the criminal justice system. Furthermore, every expert by experience was actively involved in supporting women, either through local Maternity and Neonatal Voices Partnerships, working in a perinatal mental health charity, or expert by experience roles within mental health services. This blurring of participant categories was evident throughout the study, complicating the classification process. These overlapping participant categories underscore the complexity of trauma discussions and highlight the need for sensitivity and flexibility in research classification.

### Overview of findings

Participants offered a range of insightful perspectives about trauma discussions which have been summarised in five themes. The first, ‘Rationale for discussions’, explores whether care providers should raise the issue of previous trauma with women. ‘Professionals and settings’ considers which professionals should carry out trauma discussions and the optimum setting for these conversations. The third theme, ‘Timing considerations’, examines when trauma discussions should be carried out. In the fourth theme, ‘Communicating about trauma’, interviewee perspectives on discussing trauma are described. The final theme, ‘Supporting care providers’, addresses the training and emotional well-being needs of professionals conducting trauma discussions. Healthcare professional participants were designated as ’HP1’, ‘HP2,’ and so forth, while individuals with lived experience of trauma were coded as ’WLE1’, ‘WLE2,’ and experts from the voluntary sector as ’EV1’, ‘EV2,’ and so on.

### Rationale for discussions

Participants highlighted the profound impact of sensitively addressing trauma and providing post-disclosure support in the perinatal period, with one interviewee describing it as *‘an amazing opportunity and time to do this critical work’* (HP5). We heard from participants that these discussions offered an opportunity for women to reclaim agency over their prior experiences, rather than erasing traumatic memories. They highlighted the interconnectedness of mental health and trauma experiences, suggesting that discussions surrounding these topics should be integrated. Some participants suggested a devastating link between trauma and suicide, arguing that without trauma discussions, care providers miss the opportunity to support women who may be extremely distressed. One expert by experience candidly expressed the potentially transformative impact of trauma discussions and support, commenting:

> *‘I think it [talking about previous trauma and providing support] can make the difference between, it sounds dramatic but life and death. Literally. After my first birth I thought about ending my own life and this time I obviously don’t anticipate that happening’* (WLE1).

Care providers were seen as having an important role to play in educating women about the effects of trauma and preparing them for the possibility that the perinatal period might be challenging. Simply having the opportunity to talk was viewed as beneficial to women: *‘they feel lighter, they feel like they share their burden, they feel like they can get better’* (EV4), and had the potential to greatly improve children’s lives, *‘interrupting that intergenerational transmission of trauma’* (HP5). Participants suggested that care providers should support both parents, with some proposing that partners should also be asked about previous trauma and mental health. Interviewees also underscored the potential economic benefits of implementing properly funded trauma discussions. A psychiatrist participant argued that early trauma discussions are a good investment in time to pick up problems early and ensure women are given the support they need, pointing out *‘that is better for her, but it is also actually a more efficient way to run the service’* (HP3).

However, interviewees cautioned that trauma discussions will only be of value to women if they are used to improve care, rather than merely *‘just asked, recorded, then ignored…’ (EV7).* Broader support services, particularly mental health services, were viewed by some as inadequate, inconsistent, or not trauma-informed. One healthcare professional participant cautioned that insensitive trauma conversations could be distressing and cause women to confront past experiences in an unanticipated and harmful way: *‘that may not have been a big deal to them then all of a sudden oh my god that was abuse’* (HP1). Concerns were also raised about overzealous safeguarding responses to trauma disclosures, with potentially catastrophic consequences:

> *‘A good outcome is that that woman has a more positive experience of being pregnant and giving birth and the early time with her child, than she would have done without us asking. If the reality is that only 1 of 10 women who we ask has that outcome and 9 of them have disasters because suddenly social services are involved, and they are beholden to all sorts of systems and they are reporting to the police and they didn’t really want to…’ (EV1)*.

### Professionals and settings

When exploring who should conduct trauma discussions, most experts by experience and voluntary sector practitioners felt that women would be most comfortable disclosing previous trauma to a female clinician, with some explicitly stating they would not disclose previous trauma to a male: *‘if it was me I would lie I wouldn’t even open up to a man’ (EV4).* While it was felt that any maternity care provider could potentially initiate trauma discussions, midwives and health visitors were particularly favoured due to their frequent contact with women during pregnancy and the postnatal period. Continuity of care was also perceived as important, enabling professionals to build rapport and create a psychologically safe environment for discussions:

> *‘It is such a personal and very intimate part of your life, it is not something you are ready to share with a complete stranger who says ‘oh hello I am your midwife, now tell me have you ever experienced trauma?’ (*EV6).

Nonetheless, some participants noted that even where continuity is not possible, care providers can use kindness, compassion, and warmth to create a psychologically safe environment.

Many participants expressed concerns about discussing trauma in the clinical setting, due to its potential to inhibit disclosures. A perinatal mental health specialist midwife vividly described the lack of privacy in many clinical environments, saying her antenatal clinic was *‘like Grand Central Station’* (HP2). Participants suggested that a more informal environment, with comfortable seating and refreshments, would be more conducive to sensitive discussions. For some women, clinical environments were reminiscent of previous negative experiences with statutory services. It was suggested that there should be support available for women who become upset and need space to collect themselves after the appointment, in terms of both a private space and a staff member.

### Timing considerations

Participants stressed the need for maternity care providers to initiate conversations about previous difficult experiences only when sufficient time is available to listen and respond effectively. One participant cautioned:

> *‘If you have got 2 minutes left and you say to somebody ‘so have you ever experienced sexual trauma?’, no, just don’t do it. Do it on a different occasion, think practically about it. Have you got the time to give the space?’* (HP10).

It was unanimously agreed that discussions about trauma should not occur in front of partners or young children, as their presence could inhibit open discussion. Participants also highlighted the importance of forewarning women about upcoming discussions on previous trauma to allow them to prepare and arrange for support if needed. Participants noted that for many women, multiple encounters are necessary before they feel safe enough to share their histories, with one emphasising the *‘enormous amount of weighing up that will go on before people trust and disclose’* (HP9). Some participants proposed that even where women choose not to disclose on that particular occasion, carrying out routine trauma discussions sensitively could facilitate trust and future disclosure: *‘you have planted the seed of if I am ever ready I can. This is a safe space. This is a safe person’* (WLE2).

Participants acknowledged the challenges associated with discussing trauma during the first midwifery appointment due to time constraints and the predominantly closed-ended format of the appointment. One participant expressed her unease at having to move on from an emotive disclosure to *‘do you have a dentist, do you have a dog kind of thing’* (HP11). Participants from all population groups felt that an additional antenatal appointment specifically focused on emotional health and well-being, including discussions about previous trauma would be helpful. Comments included*, ‘I just got goosebumps just thinking how good that would be. Yes’ (EV4); ‘I think that’s brilliant’* (WLE4), and *‘I think it would be wonderful. And I think it would really do a lot to allay fears of women’* (EV6). A health advocate described it as a *‘great idea’* (EV5) and added: *‘even things that we don’t share with our husbands will come out, our worries, our fears.’* They highlighted advantages such as alleviating the crowded schedule of the first maternity care appointment and providing a protected space for meaningful conversations. Participants favoured an unstructured, woman-led conversation format, with one remarking, *‘no paperwork, you just go along and you hear and connect. That is really powerful’* (EV3).

### Communicating about trauma

Participants highlighted the complexities of discussing trauma, noting the need for sensitivity, clarity, and accuracy. They noted that commonly used terms such as ‘trauma’, ‘emotional abuse’, ‘sexual abuse,’ and ‘physical abuse’ may not resonate with women’s own perceptions of their experiences, potentially hindering disclosure. For instance, one participant explained that women may not feel they have been abused *‘but if you knew her history you would think she absolutely was’* (EV2). Some groups of women, such as those who are autistic or have learning disabilities, were felt to face additional challenges in understanding and articulating their experiences. Further, participants underscored the importance of developing materials with low literacy levels in mind. The use of explicit or formal language was perceived to inhibit conversations, potentially causing mothers to *‘completely shut off’* (HP4). It was felt that closed-ended questions in general may deter women from disclosing because of a fear of social services involvement.

Participants universally felt that communication challenges are magnified for women with limited English proficiency, who may struggle to grasp complex information or nuances, leading to misunderstandings or embarrassment. Participants highlighted simply translating questionnaires into a woman’s first language does not guarantee her understanding as not all women are literate in their native language. The stigma surrounding mental health in some cultures can further hinder open discussions; a midwife shared her experience of women reacting with discomfort to mental health enquiries: *’the woman will look at the partner or the granny as if to say, ‘this is awful that you are even asking me this’* (HP11).

Many participants questioned the effectiveness of quantitative trauma and mental health screening tools, advocating for a relational approach instead. Views on specific screening tools varied. While some considered the Antenatal Psychosocial Risk Questionnaire (ANRQ) to be clear and comprehensive, others questioned the utility of detailed questions such as ‘when you were growing up, did you feel your mother was emotionally supportive of you?’ in the absence of clear pathways for intervention or support. Some found the direct nature of the ANRQ questions intrusive and *‘more like child protection, you are looking if I am going to be a good mum’* (WLE3). The Kimberley Mum’s Mood Scale was widely praised for its simplicity, sensitivity, and visual approach, which participants felt fostered trust and honest responses. One interviewee remarked, *‘it is simple but very, very effective’*. In contrast, the Adverse Childhood Experiences questionnaire (ACE-10) faced strong criticism for its explicit language and potential to re-traumatise women. An expert by experience expressed a visceral emotional reaction to the questionnaire:

> *‘It reminds you that you weren’t looked after. You weren’t taken care of. You know that as a child, you weren’t parented, you weren’t loved in the way that a child should be loved. What upsets me isn’t the act of the abuse, it is the fact that I wasn’t looked after and I didn’t have that love and care and what that means as an adult’* (WLE2).

Participants stressed the need for sensitive communication when women disclose previous trauma, advocating for active listening over intrusive questions: *‘understand the difference between your own nosiness versus what is actually needed’* (HP10). Participants stressed the importance of providing independent access to support for women who choose not to disclose previous trauma. They also highlighted the need for care providers to adopt a universal precautions approach, being sensitive to the possibility of trauma in every interaction:

> *‘Treating everybody with respect and coming from a place of actually you could have had a really horrible story…’* (WLE1).

### Supporting care providers

Participants felt that all staff, including receptionists and clinical support, should be trained to recognise signs of possible trauma and communicate these observations to maternity care providers. Interviewees stressed the importance of recognising non-verbal cues indicating trauma or mental health struggles, with some experts by experience expressing frustration at care providers’ failure to pick up on their distress, for example *‘it was quite clear that I was distressed but they just didn’t seem to realise’* (WLE1). Interpersonal skills, centred on compassion and relationship-building, were deemed essential. although challenging to teach. Simulation with actors was suggested to enhance communication skills. Participants also considered that training in fundamental counselling skills could aid in supporting women upset during discussions of previous trauma.

Multiple interviewees talked of the burden of hearing trauma disclosures with one describing it as *‘heavy going’* (HP4), and proposed that these conversations may be particularly poignant for care providers who have endured similar experiences themselves. Participants suggested that awareness of the potential for hearing upsetting stories could mean care providers are reluctant to carry out discussions about previous trauma. Moreover, participants cautioned against the potential devastating consequences of interruptions and premature termination of conversations caused by provider discomfort:

> *‘I spend so much of my time as a psychosexual therapist unpicking how clients have felt about being shut down by healthcare professionals, because they have been asked a question, but they haven’t been heard and listened to […] It is very likely to be about time restrictions, or it has triggered something in this professional. But your client shouldn’t have to carry that, your client just goes, oh I will never tell them again.’* (HP10).

Participants emphasised the essential role of supportive management, both in managing caseloads to ensure that emotionally challenging work is distributed evenly among the team, and recognising and supporting staff who are suffering due to their own difficult life experiences. However, interviewees expressed concerns that staff might not feel comfortable disclosing trauma or subsequent mental health struggles to management due to fears of career repercussions. Further, managers themselves voiced concerns that staff support services could be seen as punitive rather than supportive. A therapist participant criticised the prevailing culture within the NHS that discourages vulnerability and prioritises stoicism, stating,

> *‘Not wanting to be seen as weak or not able to cope, this ideology which is really strong in healthcare that you have just got to crack on with it, come on this is the job, pull up your pants, this is what you signed up for. It is not helpful, and it stops people from disclosing when the shit is hitting the fan for them’* (HP10).

The consensus among participants was that staff expected to engage in routine trauma discussions should receive regular reflexive supervision during working hours and from someone independent of the maternity team. Both group and individual supervision were deemed valuable and complementary. An expert by experience denounced the expectation for staff to conduct these discussions without proper supervision as *‘completely unfair and inappropriate’* (WLE4). Stressing the importance of mandatory supervision, a therapist noted that without it, staff may not recognise the potential for burnout or seek support, stating, *‘you don’t know until you know how beneficial it is’* (HP10).

## Discussion

While participants argued that it is crucial for maternity care providers to raise the issue of previous trauma, significant logistical challenges were highlighted. Interviewees noted that discussing trauma requires care, respect, multi-cultural sensitivities, time, and vulnerability. The study found that discussing previous trauma is a complex intervention that requires careful consideration of methodology, setting, timing, referral pathways, communication with women, and staff training and support. Drawing upon insights from the interviews and findings from a previous systematic review and qualitative synthesis (Cull et al., 2023), a framework of guiding principles has been developed to provide practical recommendations for healthcare professionals. The framework aims to navigate the challenges associated with addressing previous trauma effectively. The development and evaluation of this framework will be discussed in a subsequent paper. Interviewees indicated that discussing previous trauma can trigger distress, including feelings of shame, embarrassment, and painful memories. Despite the increasing use of quantitative trauma measurement tools such as the Adverse Childhood Experiences questionnaire (Felitti et al., 1998) in healthcare, including maternity care, concerns persist, as identified in this study, about their limitations and potential harms (Anda, Porter & Brown, 2020; Lacey and Minnis, 2020; Walsh, 2020). Pregnant women may fear the impact of their trauma on their unborn child, leading to heightened anxiety and a sense of disempowerment. Gentry and Paterson (2021) argue against routine ACE screening due to a lack of effective interventions for those with positive ACE scores and uncertainty about the balance of benefits and harms. Similarly, Finkelhor (2018) criticises ACE screening for its ineffectiveness in identifying and addressing the needs of individuals affected by previous trauma. The Kimberley Mum’s Mood Scale, which incorporates a visual Likert scale derived from the Edinburgh Postnatal Depression Scale alongside a discussion of key well-being domains including childhood experiences and mental health, emerged as the preferred tool among the interview participants (Marley et al., 2017; Cox, Holden, and Sagovsky, 1987). While participants felt the scale was likely to be acceptable to women and effective in encouraging open conversations, its design for Aboriginal women in Western Australia necessitates substantial adaptation for use elsewhere. Consequently, there is a pressing need for a culturally sensitive, co-designed tool for use in maternity services.

The study highlights a key challenge in current maternity care practices: broaching the topic of prior trauma during the initial booking appointment often proves ineffective and may distress women who are unprepared for such discussions. This finding, supported by feedback from interview participants, underscores the need for a separate antenatal appointment specifically focused on emotional health and well-being. This approach would provide multiple opportunities for women to address trauma-related concerns and mental health, facilitating more open and sensitive discussions. Moreover, continuity of carer was emphasised as crucial for building the trust necessary for these sensitive conversations. However, achieving this within the constraints of overstretched and understaffed maternity services, which are increasingly driven by performance standards and have failed to invest in preventative care, remains a significant challenge (Darzi, 2024). Addressing these barriers will require systemic changes, including staff training, resource allocation, and a reorganisation of appointment structures to prioritise emotional well-being as part of routine care.

The study explores the emotional challenge care providers face when hearing disclosures of previous trauma, especially for those who have experienced trauma themselves. Survivors of trauma may find that supporting women who have faced similar experiences can evoke distressing memories (Donovan et al., 2021). It is essential that staff involved in trauma discussions be provided with independent, professional support services. Staff may be reluctant to share personal experiences with colleagues they know or may not recognise when their stress and burnout levels are escalating. A fundamental culture shift is necessary to ensure staff access available support. This transition necessitates moving beyond merely ’offering’ support to those in need, towards integrating support as a routine aspect of daily work life, actively provided to all staff during working hours. Such a cultural transformation has the potential to significantly improve the effectiveness of support initiatives (Clarkson et al., 2023).

### Strengths and limitations of the study

There is a lack of clear guidance on the practical implementation of trauma-informed perinatal care, which the study addresses. One of its key strengths lies in the engagement of a diverse range of stakeholders, including the Research Collective who supported the study throughout. Further, the interview sample included various professional groups involved in delivering maternity care, diverse trauma types and experts by experience from marginalised groups. This collaborative and broad-ranging perspective bolsters the validity of the findings and their relevance to a range of settings.

While the study provides valuable insights into conducting effective and sensitive trauma discussions, several limitations affect the interpretation of the findings. The use of purposive sampling potentially introduced selection bias. The decision to conclude interview recruitment after 23 participants, driven by data richness and manageability, may have excluded diverse perspectives that additional participants could have brought. Despite efforts, challenges in recruiting women with limited English proficiency persisted. Further, the absence of participants from Asian backgrounds is notable, given their heightened risk of poor maternal and neonatal outcomes (Knight et al., 2020). Future research should prioritise the inclusion of this demographic group and other underrepresented groups.

## Conclusion

This study presents findings from semi-structured interviews with a range of expert stakeholders. Key findings include the importance of time, adequate support for staff, and effective referral mechanisms to enable maternity care providers to initiate discussions about previous trauma with women, and to ensure appropriate personalised follow up in the event of disclosure. Central to these conversations are trust and relationships, necessitating careful consideration in care provision. Furthermore, the research underscores the necessity of comprehensive staff training and support.

The interview findings illuminate critical aspects of trauma-informed perinatal care, offering valuable insights into the complexities and challenges faced by both women and maternity care providers. By focusing on routine trauma discussions during the perinatal period, the research addresses a significant gap in existing literature and provides practical solutions to improve the quality of care for women who have experienced trauma.

## Data Availability

All data produced in the present study are available upon reasonable request to the author.

## Acknowledgements

We thank the EMPATHY study Research Collective for their invaluable contributions to this study: Laura Abbott, Juliet Albert, Kirsty Armstrong, Jill Benjoya Miller, Ang Broadbridge, Emma Brooks, Geraldine Butcher, Jo Doherty, Amber Jackson, Isobel Martin, Elsa Montgomery, Sam Pointon, Sarah-Jayne Pomeroy, Erjola Sadria, Gill Skene, Memuna Sowe, Kim Thomas, and Lucy Warwick-Guasp.

## Appendix 1 Topic guide for all interviewees

### HOW SHOULD MATERNITY CARE PROVIDERS ASK ABOUT DIFFICULT PAST EXPERIENCES?

1. Do you think maternity care providers **should** ask pregnant women about difficult past experiences?
2. **When** should maternity care providers ask about difficult experiences? PROMPTS:

- At booking?
- At a later routine appointment?
- At a separate appointment for this purpose?
- At multiple appointments?
- Is continuity of care / existing relationship important?
3. **Who** should ask about difficult experiences? PROMPTS:

- Is professional background important (midwife / care assistant / obstetrician?)
- Is the gender of the person asking important?
4. **Where** should these discussions take place? PROMPTS:

- Is this important?
- Home / clinic - hospital or community setting?
- What if partners / children are present?
5. How can maternity care providers ensure that women who want to discuss their histories **feel comfortable** to do so? PROMPTS:

- How can the questions be asked sensitively?
- Should partners be excluded from part or all of an appointment to allow these issues to be discussed in private?
- For women who want to disclose their histories, what do you think would prevent them from doing so?
6. **How** should maternity care providers ask about difficult past experiences? PROMPTS:

- How should the question be asked - direct question? General discussion? Questionnaire?
- Show examples:

▪ Antenatal Psychosocial Risk Questionnaire (ANRQ) (Austin et al., 2013)
▪ ACE-10 Questionnaire (Felitti et al., 1998)
▪ Kimberley Mum’s Mood Scale (Marley et al., 2017)
▪ Trauma History Questionnaire (Green, 1996)
▪ Hypothetical Prompt developed by White, Danis and Gillece (2016)
▪ Prompt developed by Montgomery (does not explicitly name abuse) ‘Sometimes pregnancy can trigger unexpected memories of things that have happened to you or feelings that can take you by surprise. If that happens to you and you would like to talk about it, please let me know’.
- Complete through conversation with maternity care provider, or self-complete (on ipad or paper) then discuss with maternity care provider?
- Another option is to complete independently from maternity care, e.g. online tool which encourages women to seek support from healthcare provider but also provides links to relevant third sector agencies
- How can the questions be asked sensitively?
7. How should maternity care providers **prepare women** for this conversation and let them know the purpose of the discussion? PROMPTS:

- What are the issues around confidentiality?
- Should we talk with women about potential negative implications of disclosures (for example, if children / self potentially at harm maternity care provider will need to share this information)?
- Should we talk with women about the potential positive implications of disclosure? To receive support and understanding, be offered adaptations to care, signpost or refer to services that might help in healing.
8. Are there any additional considerations when discussing trauma with **women who don’t speak English as a first language**?

- Challenges around use of interpreters
- Looking back at methods of discussing trauma with women (e.g. Kimberley Mums Mood Scale), how easy would they be to understand for women with limited English?
- Would women prefer to self-complete trauma checklist in own language?
9. Some of the **language used around difficult past experiences** may make these conversations harder for women. Are there any terms that you think maternity care providers should avoid? PROMPTS

- Examples - victim / survivor
- Trauma / abuse / difficult experiences
- Maternity care providers have said that when they’re talking about caring for women who have experienced trauma, they’re not sure how to refer to them. For example, survivor moms. Do you have any thoughts about this?

### HOW SHOULD MATERNITY CARE PROVIDERS RESPOND TO DISCLOSURES OF PREVIOUS TRAUMA?

10. How should the information be **recorded and shared**? PROMPTS:

- Within the maternity team
- With wider support services e.g. Health Visitor, GP, neonatal team, perinatal mental health, safeguarding, third sector
- Consent for this
- Limits of confidentiality
- Electronic / hand-held record
11. What **information and support** might women who have had difficult experiences find helpful? PROMPTS:

- For example mental health specialists / support from the voluntary sector
- Are there any other helpful resources you are aware of, for example books or websites?
- Do you think tailored small group antenatal classes for women who have had difficult experiences would be beneficial? (like Centering Pregnancy)
- How about peer or lay support groups?
- Or groups with more of a social focus, not explicitly about trauma but offering arts / movement / meditation?
- Using a friends and family diagram (genogram) - strengths based approaches to help women identify the sources of support in their lives and communities?
- Can you think of any services which aren’t currently offered, but would be helpful to women?
12. How can **maternity care be adapted** to help women who have had difficult experiences? PROMPTS:

- E.g. continuity of carer, limited vaginal examinations, elective caesarean section
13. What do you think should be included in **training for maternity care providers** around trauma-informed care? PROMPTS:

- Training in communication skills to sensitively ask about previous trauma and respond to disclosures?
- Instances in which safeguarding procedures will, and will not, have to be followed, and how to discuss these with women.
14. **What difference** do you think discussing prior trauma with women and providing support could make?

- What matters to women?
- What should we be measuring?
15. Is there anything I haven’t asked that you would **like to add**? PROMPT

- Do you think specific conditions need to be in place before routine discussion of trauma is introduced?
- Was it helpful to receive the topic guide in advance?

### ADDITIONAL QUESTIONS FOR HEALTHCARE PROFESSIONALS AND VOLUNTARY SECTOR EXPERTS ONLY

16. How can **adequate time be ensured** for these conversations to be meaningfully had within an overstretched and understaffed service?
17. How can women who have experienced trauma be recognised through **verbal and non-verbal signals**? PROMPTS:

- How can care providers support women who they suspect are experiencing the effects of trauma, but have chosen not to disclose?
- Some providers have talked about ‘universal precautions’: assuming all women have experienced trauma as it is so prevalent. How do you think care providers can adjust their care to avoid causing distress to women who have had difficult past experiences?
18. Trauma disclosures can be distressing to hear. How can **midwives’ emotional wellbeing be protected**? PROMPTS:

- Supervision models -
i. Who should provide the support - a midwife within the trust (Professional Midwifery Advocate) or a trained therapist?
ii. Should this be on an as-requested basis or with regular meetings (automatic supervision)?

- Midwives who have been, or are currently in, violent or abusive situations may find training around this issue distressing, and may find these conversations difficult. How can they best be supported?
- Should midwives be able to opt out of having these discussions? Some midwives may not have the personal resources to support women around these issues due to their own trauma.

